# Association between sex and race and ethnicity and intravenous sedation use in patients receiving invasive ventilation

**DOI:** 10.1101/2024.04.04.24305330

**Authors:** Sarah L. Walker, Federico Angriman, Lisa Burry, Leo Anthony Celi, Kirsten M. Fiest, Judy Gichoya, Alistair Johnson, Kuan Liu, Sangeeta Mehta, Georgiana Roman-Sarita, Laleh Seyyed-Kalantari, Thanh-Giang T. Vu, Elizabeth L. Whitlock, George Tomlinson, Christopher J. Yarnell

**Author notes:** Corresponding Author, Christopher Yarnell, Department of Critical Care Medicine, Scarborough Health Network, 3030 Lawrence Ave E, M1P2V5, Toronto, ON, Canada.

## Abstract

**Rationale:** Intravenous sedation is an important tool for managing invasively ventilated patients, yet excess sedation is harmful, and dosing could be influenced by implicit bias.

**Objective:** To measure the association between sex, race and ethnicity, and sedation practices.

**Methods:** We performed a retrospective cohort study of adults receiving invasive ventilation for 24 hours or more using the MIMIC-IV (2008-2019) database from Boston, USA. We used a repeated-measures design (4-hour time intervals) to study the association between patient sex (female, male) or race and ethnicity (Asian, Black, Hispanic, White) and sedation outcomes. Sedation outcomes included sedative use (propofol, benzodiazepine, dexmedetomidine) and minimum sedation score. We divided sedative use into five categories: no sedative given, then lowest, second, third, and highest quartiles of sedative dose. We used multilevel Bayesian proportional odds modeling to adjust for baseline and time-varying covariates and reported posterior odds ratios with 95% credible intervals [CrI].

**Results:** We studied 6,764 patients: 43% female; 3.5% Asian, 12% Black, 4.5% Hispanic and 80% white. We analyzed 116,519 4-hour intervals. Benzodiazepines were administered to 2,334 (36%) patients. Black patients received benzodiazepines less often and at lower doses than White patients (OR 0.66, CrI 0.49 to 0.92). Propofol was administered to 3,865 (57%) patients. Female patients received propofol less often and at lower doses than male patients (OR 0.72, CrI 0.61 to 0.86). Dexmedetomidine was administered to 1,439 (21%) patients, and use was largely similar across sex or race and ethnicity. As expressed by sedation scores, male patients were more sedated than female patients (OR 1.41, CrI 1.23 to 1.62), and White patients were less sedated than Black patients (OR 0.78, CrI 0.65 to 0.95).

**Conclusion:** Among patients invasively ventilated for at least 24 hours, intravenous sedation and attained sedation levels varied by sex and race and ethnicity. Adherence to sedation guidelines may improve equity in sedation management for critically ill patients.

## Introduction

Intravenous sedation can facilitate invasive ventilation in critically ill patients. However, deep sedation and use of benzodiazepines instead of propofol or dexmedetomidine are associated with longer duration of ventilation and delirium.(1–3) Deep sedation is associated with delayed extubation and increased mortality.(4,5) Guidelines recommend a protocolized approach where sedation dose is titrated to light sedation using a standardized scale.(6) Implementation of this approach is not universal, and patients are often too deeply sedated.(7–10)

One potential explanation for over-sedation could be that practice varies according to patient sex or race and ethnicity due to implicit bias.(11,12) Hispanic compared to non-Hispanic clinical trial participants with moderate-to-severe acute respiratory distress syndrome (ARDS) were more likely to be deeply sedated (odds ratio 4.98).(13) Hispanic patients with COVID-19 receiving invasive ventilation were also more likely to receive benzodiazepines than non-Hispanic White or non-Hispanic Black patients.(14) In a cohort of patients receiving invasive ventilation, Black compared to White patients had an increased risk of deep sedation in the first 48 hours.(15) Fewer studies have assessed differences in sedation practices between male and female patients.

To further investigate the association between sex, race and ethnicity, and intravenous sedation practices, we performed a retrospective cohort study of patients who received at least 24 hours of invasive ventilation. We hypothesized that sedation practices would differ by sex and by race and ethnicity, such that marginalized groups receive higher doses of sedation.

## Methods

### Study Setting and Patients

We used deidentified patient data from the Medical Information Mart for Intensive Care version IV (MIMIC-IV) database.(16,17) It contains 73,181 intensive care unit (ICU) admissions from an academic quaternary centre in Boston, USA, between 2008 and 2019. It is approved for database research by the Beth Israel Deaconess Medical Center (2001-P-001699/14) and the Massachusetts Institute of Technology (0403000206).(18)

We included patients who received 24 or more hours of invasive ventilation, to focus on patients more likely to have intravenous sedation exposure, and to exclude patients admitted to the ICU post-operatively who were promptly extubated. We excluded patients with a tracheostomy noted in the first 7 days of ICU admission, which could affect the use of intravenous sedation. Patients also were excluded if they did not have a documented sex, or if their race or ethnicity was documented as “Other”, “Unknown”, or “Mixed.” We made these exclusions because the composition of these excluded groups was unknown, making the external validity of results from those groups difficult to interpret. We excluded patients categorized as “Native American / Pacific Islander” because their numbers were too few. For patients with multiple eligible ICU admissions, we included only the first.

### Design

We performed a longitudinal repeated-measures study using sedation dose per kilogram received within each time interval as the primary outcome.(19) We divided time into 4-hour intervals and followed patients from 24 hours after the onset of invasive ventilation until the earliest of 8 days from the onset of invasive ventilation, death, ICU discharge, or discontinuation of invasive ventilation.

### Variables

The primary exposures were (1) sex and (2) race and ethnicity (grouped as Asian, Black, Hispanic, and White). We considered interactions between exposures in a secondary model.

We used clinical expertise and prior research to select relevant potentially confounding variables for inclusion in the model (Figure S1). Baseline covariates included patient age, English proficiency, insurance type (Medicaid, Medicare, other); comorbidities ascertained from discharge diagnoses (see Supplement) including dementia, substance use disorder (including alcohol), and traumatic brain injury; type of ICU, year of admission; and amount of propofol and intravenous benzodiazepine administered in the first 24h of ventilation.(4)

We included time-varying covariates relevant to future sedation administration to account for time-varying severity: respiratory rate, inspired oxygen fraction, peripheral oxygen saturation, and sedation level as quantified by the Riker Sedation Agitation Scale (SAS).(20) We also included the time since ventilation initiation, and the dose of the following medications given within each time interval: vasopressors, intravenous opioids, and neuromuscular blockers (yes/no). We converted vasopressors to norepinephrine equivalents and opioids to morphine equivalents.(21,22) We built separate models for propofol, benzodiazepines, and dexmedetomidine. Each model was adjusted for time-varying use of the other two sedating agents.

The time-varying covariate values used to predict the outcome in a given interval were drawn from the previous interval, in order to ensure that covariates always preceded outcomes. For each time-varying covariate, we aggregated values measured within the same time interval using the maximum or minimum value, chosen by variable to reflect the highest severity.

### Outcomes

Our co-primary outcomes were the receipt and weight-based dose of (1) intravenous benzodiazepine and (2) propofol in each time interval. We converted benzodiazepines to lorazepam equivalents.(23) Each sedative dose was classified into one of five ordinal categories: none, or one of four quartiles of dose in cases where the dose was non-zero (Tables 2a and 2b). Secondary outcomes were dexmedetomidine receipt (yes/no) and the minimum SAS (categorized as 1, 2, 3, or ≥4) in each time interval.

### Analysis

To analyze the dose category of benzodiazepine and propofol, we used a multilevel proportional odds model, clustering by patient stay.(24) This allowed the analysis to incorporate both the probability of receiving a sedative, and the amount of sedative given, conditional on receipt. This approach used more information than a binary analysis of receipt of sedative, and had less selection bias than an analysis restricted to those who received a sedative.

For all models, we used skeptical prior distributions (normal with mean 0 and standard deviation 0.3) for fixed effects. These priors reflected our belief that large associations between a single coefficient and the outcome were unlikely.

We performed multilevel logistic regression for the dexmedetomidine outcome, clustering by patient stay. For the minimum SAS model, we only analyzed time periods where the patient was not receiving a neuromuscular blocker and had a recorded SAS value. We used a multilevel proportional odds model clustering by patient. We did not incorporate time-varying medications other than vasopressors, because we deemed those variables to be on the causal path between exposure and outcome.

The model was implemented in R 4.2.1 and parallelized on the Niagara computing cluster.(25) We summarized the posterior odds ratios (OR) using the mean and 95% credible intervals (CrI).

### Missing data

We used multiple imputation to address baseline and time-varying predictors missing in the first time interval for each patient.(26) We then used forward-fill imputation for missing time-varying covariates, similar to past analyses using this dataset.(21,27) We performed the Bayesian analysis on each dataset and then pooled the posterior distributions to capture uncertainty introduced by missing data in the final model outputs.(28)

### Sensitivity Analyses

We repeated the two primary analyses, adding interaction effects between sex and race and ethnicity. We also repeated the two primary analyses on an expanded cohort that included all patients previously excluded for having a race and ethnicity that was not Asian, Black, Hispanic, or White. This was to investigate if differences in sex persisted when including patients of unknown, other, or multiple races or ethnicities.

## Results

We studied 6,764 individuals who met the eligibility requirements (Figures S2), of whom 43% (2,924) were female; 3.5% (236) were Asian, 12% (806) were Black, 4.5% (300) were Hispanic, and 80% (5,422) were White (Tables 1a, 1b). We excluded 2,169 patients with “Other” race and ethnicity (Figure S3). Median age was 65 years (interquartile range [IQR], 53 to 76). Dementia (25%) and substance use (26%) were common.

**Table 1.**
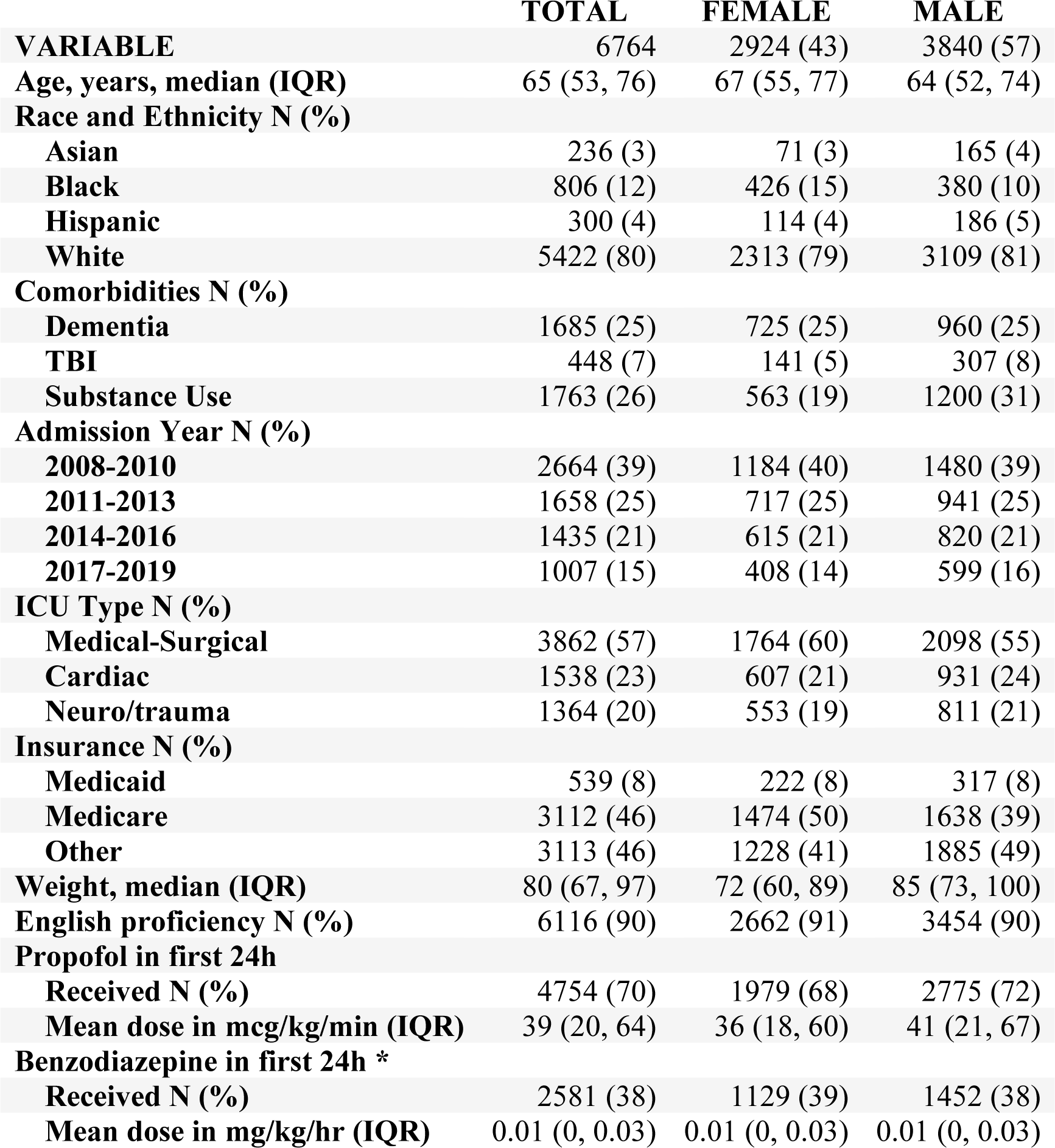
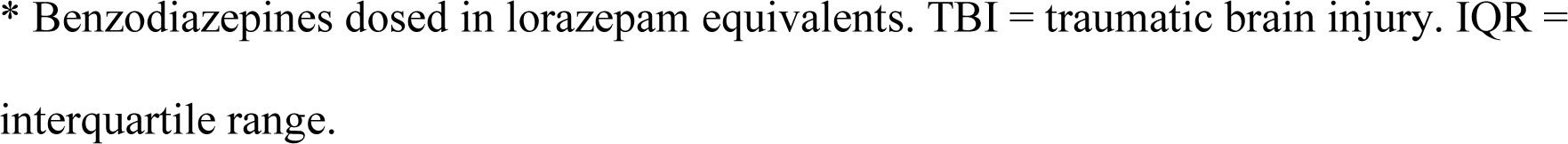
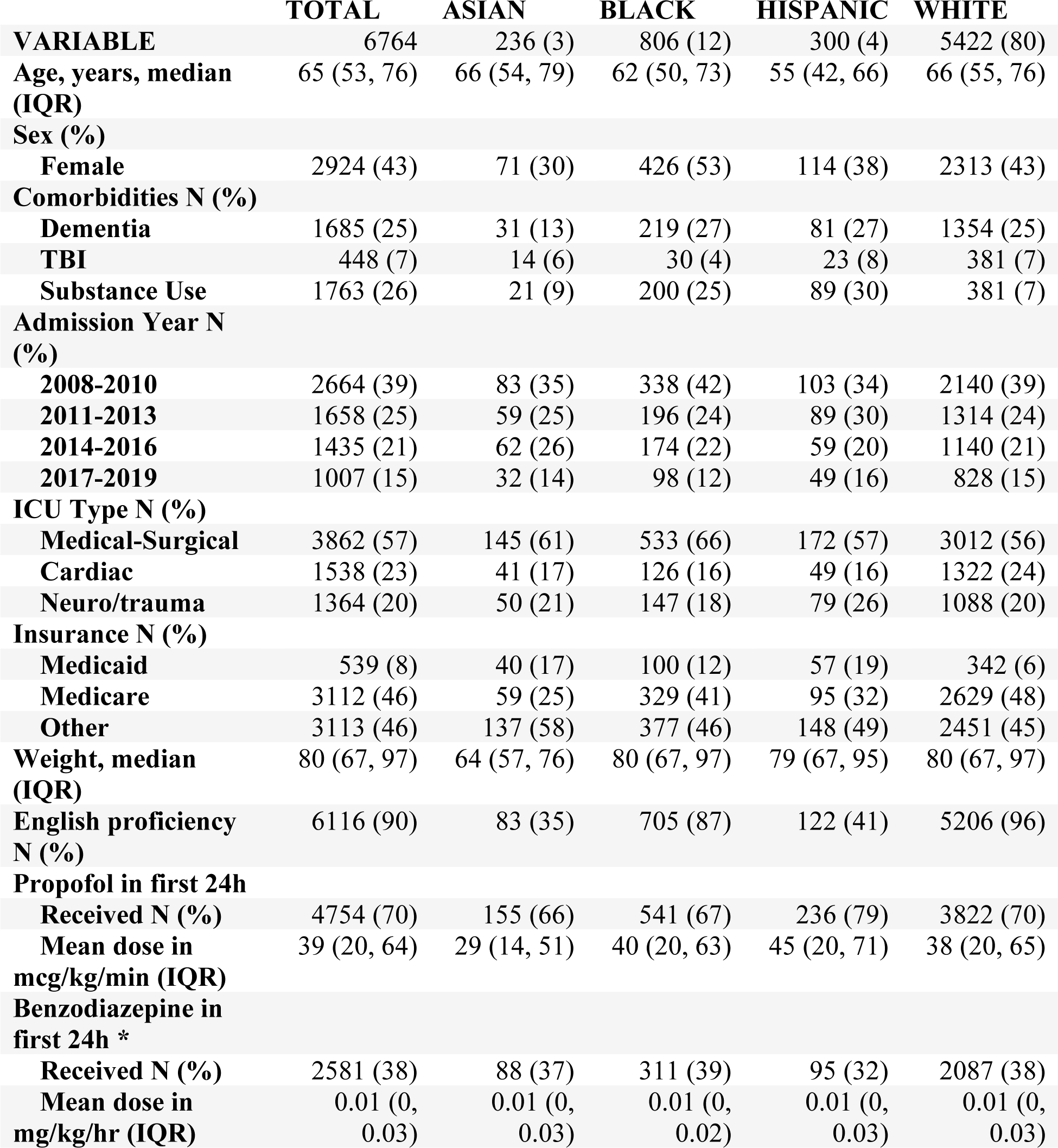
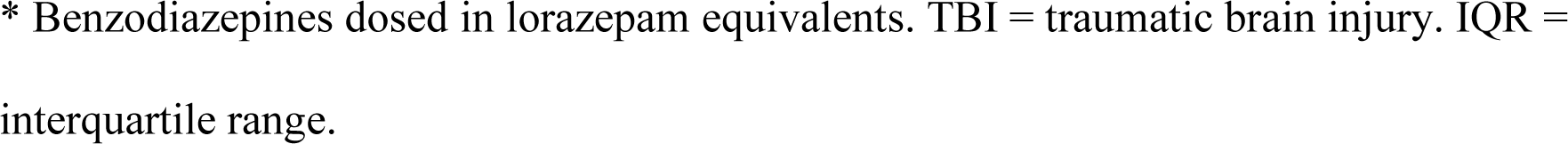
a: Baseline characteristics by patient sex b: Baseline characteristics by patient race and ethnicity

There were 116,519 intervals amounting to 466,076 hours of patient observation. Median minimum SAS was 3 (IQR 3 to 4), median respiratory rate was 22 (IQR 18 to 26), median inspired oxygen fraction was 0.4 (IQR 0.4 to 0.5), and median oxygen saturation was 97% (IQR 95% to 99%). Median duration of invasive ventilation was 3 days (IQR 2 to 6), median ICU length-of-stay was 6 days (IQR 4 to 11), and median hospital length-of-stay was 13 days (IQR 8 to 21). One-year mortality was 47%.

The variables with missing baseline data were weight (278, 4%), respiratory rate (329, 5%), inspired oxygen fraction (117, 2%), oxygen saturation (335, 5%), and minimum SAS (1,407, 21%) (Supplemental Table S3). We used 5 datasets for multiple imputation.(26,29)

### Benzodiazepines

Benzodiazepines were administered in 23% of time intervals to 2,334 (36%) patients (Table 2a) 37% of female patients and 36% of male patients received benzodiazepines. Across race and ethnicity, benzodiazepine use varied from 28% in Asian patients to 37% in White patients. In intervals where benzodiazepines were administered, median dose in lorazepam equivalents was 0.13mg/kg/hr (IQR 0.05 to 0.29).

**Table 2a:**
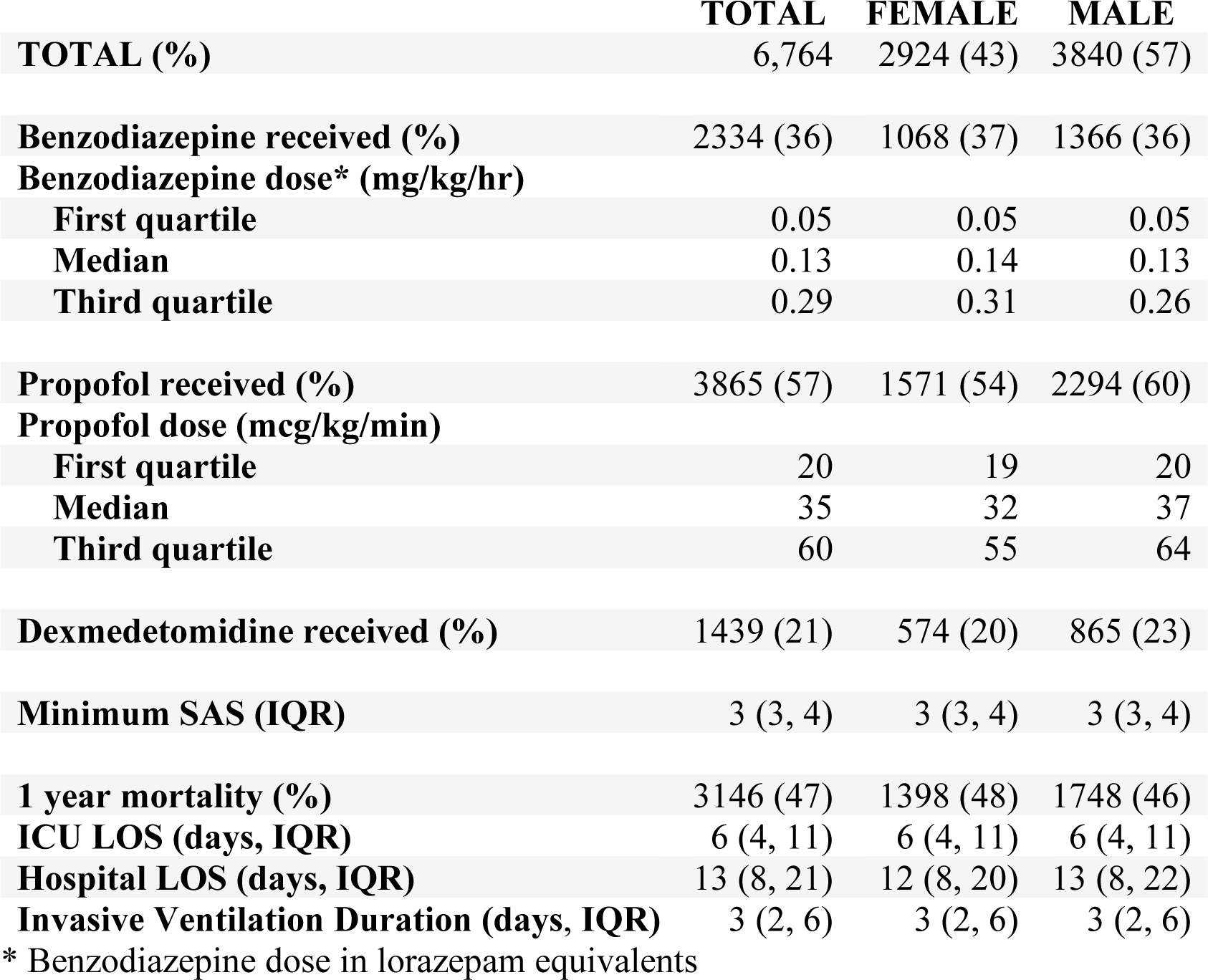
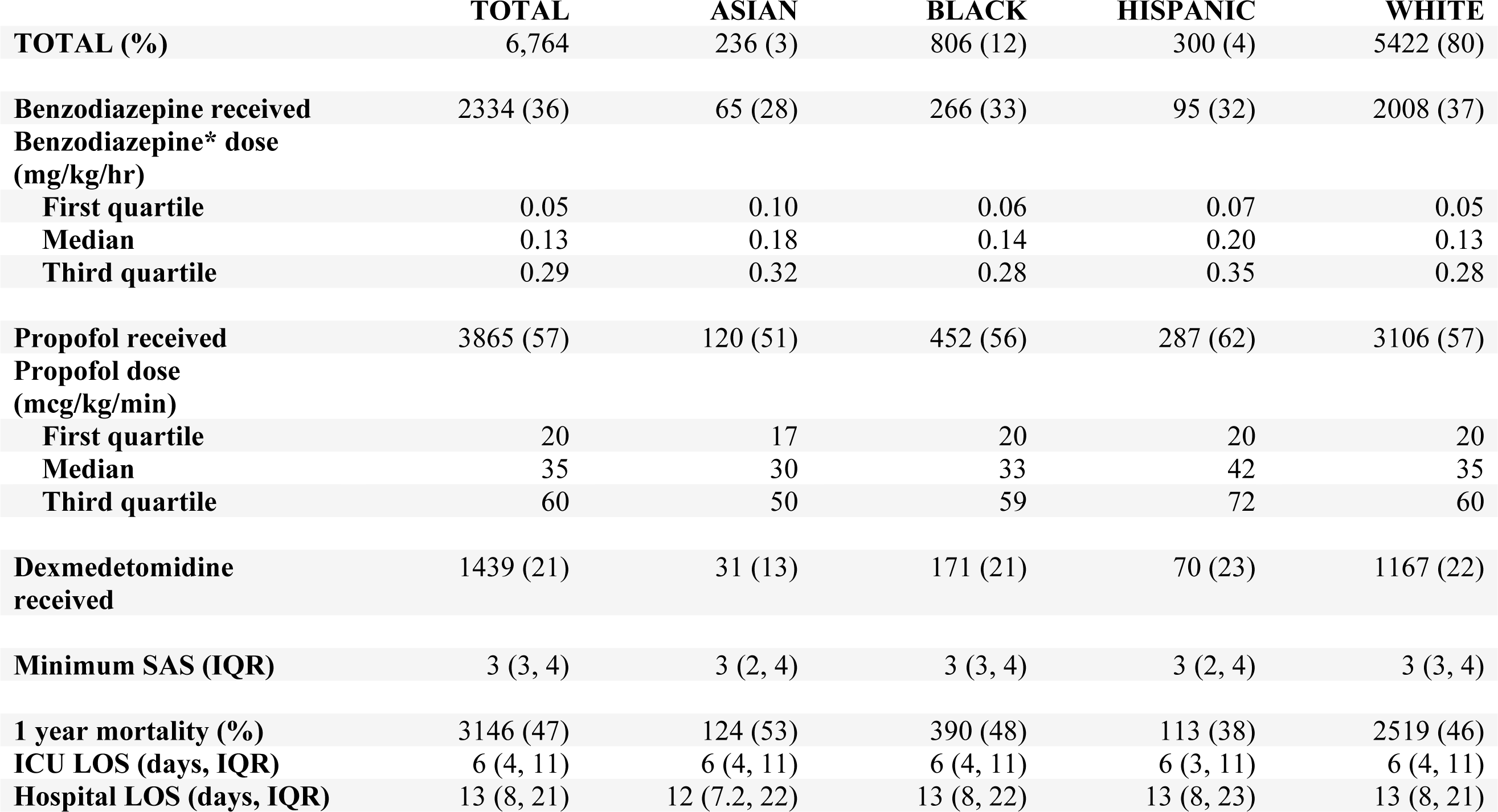
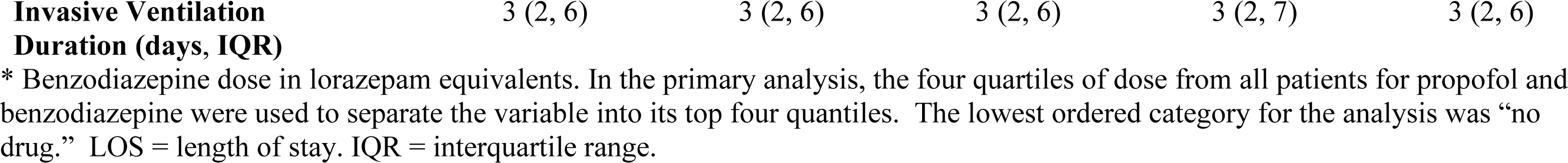
a: Sedation doses and outcomes by patient sex b: Sedation doses and outcomes by patient race and ethnicity

Female and male patients received similar doses of benzodiazepine (OR 1.09, CrI 0.89 to 1.33). Black patients received less benzodiazepine than White patients (OR 0.66, CrI 0.49 to 0.92). Using the observed 37% probability of receiving benzodiazepine for an average White patient, this translated to a 9.1% absolute reduction (CrI 1.9 to 14.6) in the probability of receiving benzodiazepine for a Black patient. Using the observed 9.3% probability of receiving more than 0.29mg/kg/hr of benzodiazepine for an average White patient, this also translated to a 2.9% absolute reduction (CrI 0.7 to 4.5) in the probability of a Black patient receiving a similar dose. The probability of receiving benzodiazepines less often and in lower doses than White patients was 96% for Asian patients and 95% for Hispanic patients (Figure 1 & S4), however, the credible interval for both odds ratios included equivalence.

**Figure 1.**
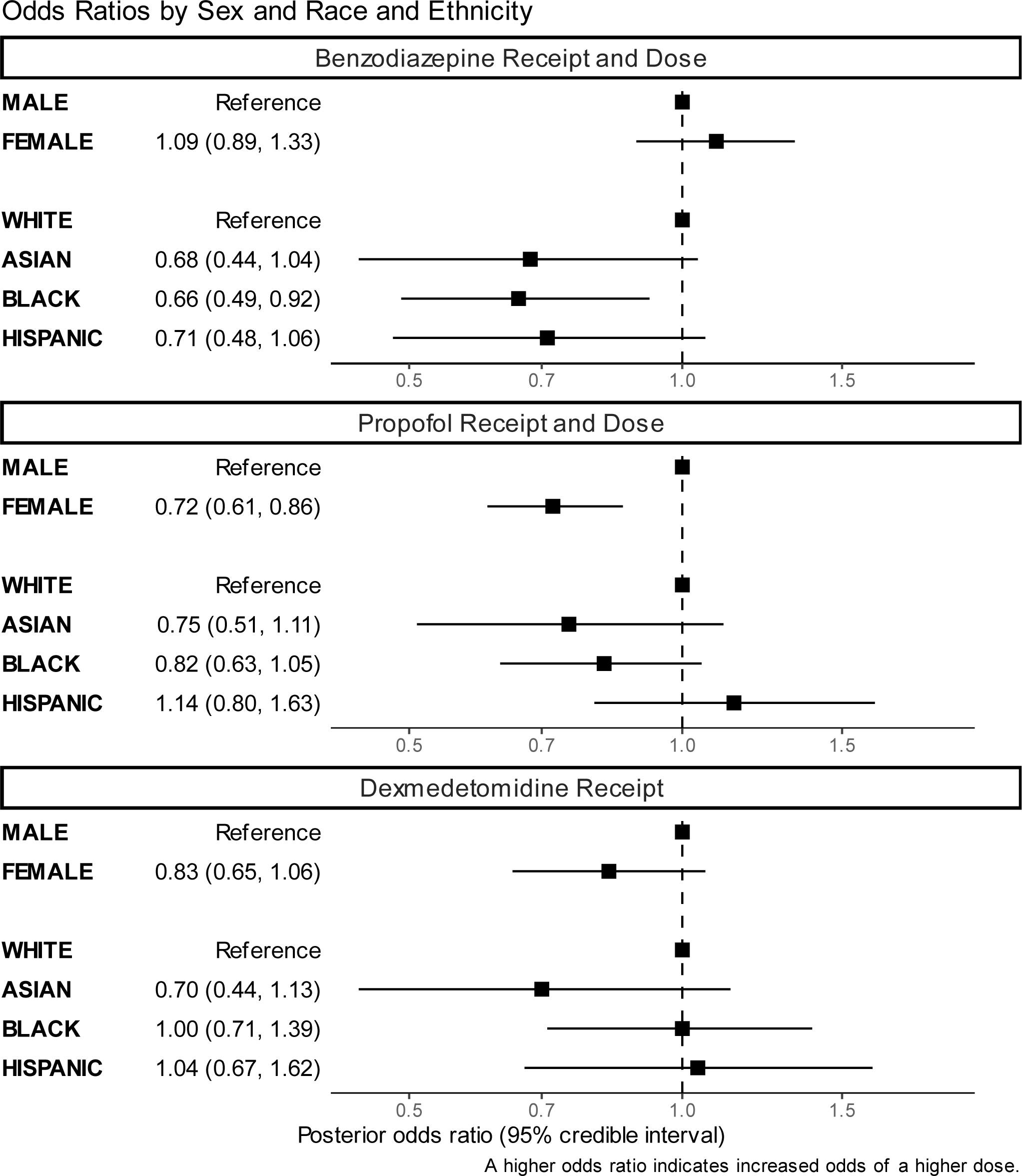
This figure shows the odds ratios for receiving a higher dose of each medication, according to patient and ethnicity, after adjusting for baseline and time-varying variables. Odds ratios less than one correspond to lower probability of receiving the medication and lower doses, while odds ratios greater than one correspond to higher probability of receiving the medication and higher doses. For dexmedetomidine, the odds ratio reflects only the probability of receiving the medication.

The sensitivity analysis including interactions showed a greater than 90% probability that male White and female White patients received more benzodiazepines than male Asian, male Black, male Hispanic, female Black, and female Hispanic patients (Figure 2 & S5). The sensitivity analysis including patients with “Other” race and ethnicity was consistent with the primary analysis (Figure S6).

**Figure 2.**
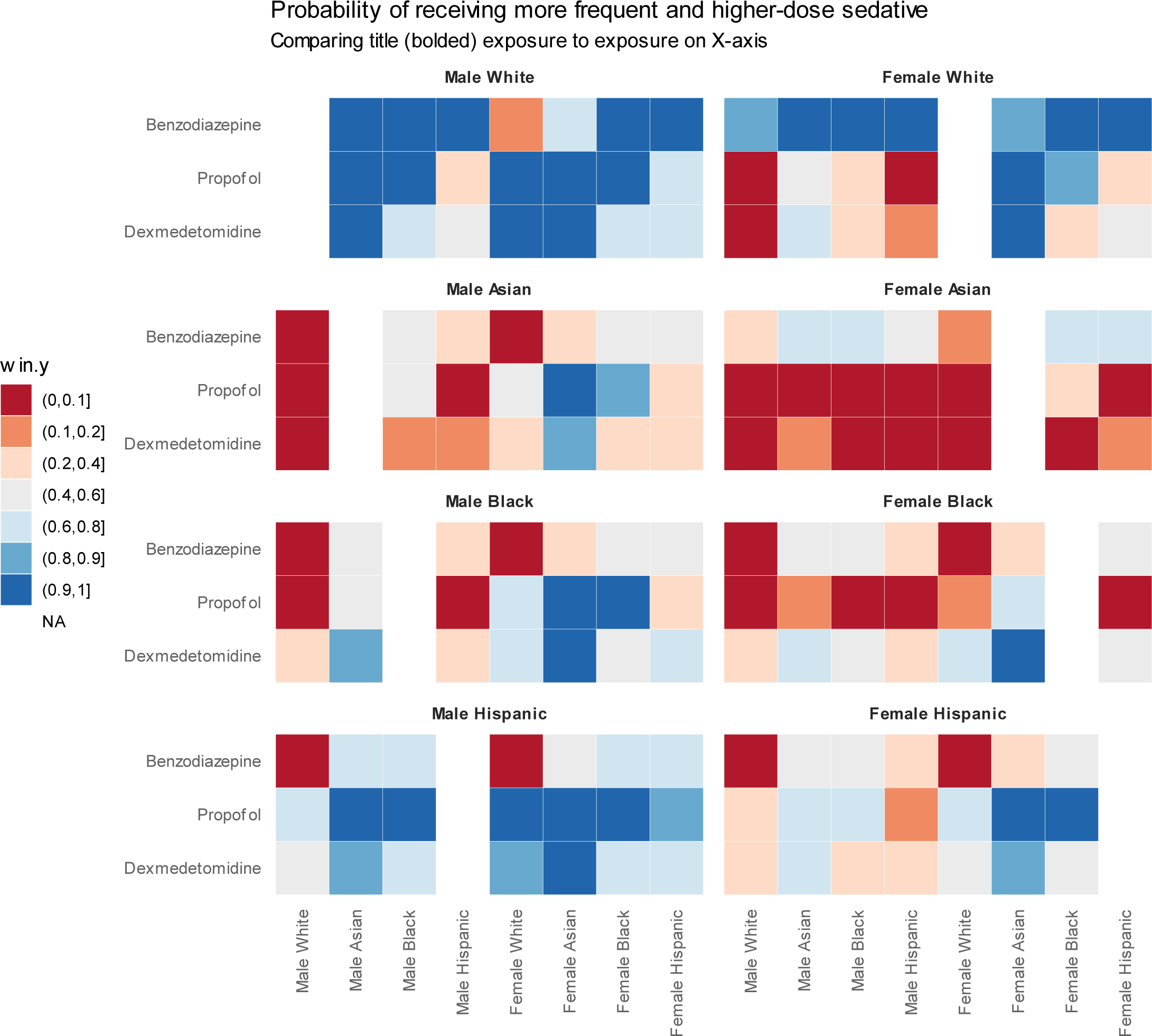
This figure compares each combination of sex and race and ethnicity (8 combinations) with respect to obability of receiving each sedative medication more often and more frequently. There are 8 tiled subplots, each corresponding to one of the 8 combinations. Each subplot has three rows, corresponding to the 3 sedative medications. The colour of each tile corresponds to the probability that the subplot’s title exposure (**bolded**) was associated with a higher odds ratio for that sedative medication than the exposure named on the x-axis. Note that there are no tiles comparing the title exposure to itself (only white space in those areas). As an example, this plot shows that male White patients had higher odds ratios than almost all other exposure combinations across the three sedatives (tiles are mostly dark blue). **If there were no differences in sedation administration by sex or race and ethnicity, then the tiles would be mostly light gray.**

### Propofol

Propofol was administered in 39% of time intervals to 3,865 (57%) patients. 54% of female patients and 60% of male patients received propofol. Across race and ethnicity, propofol use varied from 51% in Asian patients to 62% in Hispanic patients. In time intervals where propofol was administered, median dose was 35mcg/kg/min (IQR 20 to 60) (Table 2a).

Female patients received less propofol than male patients (OR 0.72, CrI 0.61 to 0.86). Using the observed 60% probability of receiving propofol for the average male or White patient, this equated to an absolute reduction in the probability of receiving propofol of 8.1% (CrI 3.7 to 12.2) for a female patient. Similarly, the probability of receiving propofol at more than 60mcg/kg/min, which was 10% in the average White or male patient, was reduced by 2.6% (CrI 1.3 to 3.7) for a female patient. The probability of receiving propofol less often and in lower doses than White patients was 93% for Asian patients and 94% for Black patients, but the credible interval of the odds ratio included equivalence. Propofol administration was similar between Hispanic and White patients (Figure 1 & S7).

The sensitivity analysis including interactions showed a greater than 90% probability that male White and male Hispanic patients received more propofol than male Asian, male Black, female White, female Asian, and female Black patients (Figure 2 & S5). The sensitivity analysis including patients with “Other” race and ethnicity was consistent with the primary analysis (Figure S6).

### Dexmedetomidine

Dexmedetomidine was administered in 10% of time intervals to 1,439 (21%) patients. 20% of female patients and 23% of male patients received dexmedetomidine. Across race and ethnicity, dexmedetomidine use varied from 13% in Asian patients to 23% in Hispanic patients. (Table 2a)

All odds ratios in the analysis without interactions had credible intervals overlapping 1. However, male patients probably received more dexmedetomidine than female patients (probability 93%), and White patients probably received more than Asian patients (probability 93%) (Figure 1). The sensitivity analysis with interactions showed a greater than 90% probability that female Asian patients received less dexmedetomidine than White and Black patients of both sexes and male Hispanic patients; for the remaining two combinations of sex and race and ethnicity (female Hispanic, male Asian) there was an 80-90% probability that female Asian patients received less dexmedetomidine (Figure 2 & S8). In the sensitivity analysis including patients with race and ethnicity classified as “Other”, it was more certain that female patients received lower doses compared to men (probability > 99%) (Figure S6).

### Attained sedation level

SAS was recorded in 102,964 (88%) intervals across 6,311 (93%) patients. Across all intervals, the minimum SAS was 1 for 9,361 (9%), 2 for 12,764 (12%), 3 for 36,506 (36%), and 4 or more for 44,333 (43%). Of the intervals with minimum SAS of 4 or greater, the minimum SAS was 5 for 2,644 (3%), 6 for 445 (0.4%), and 7 for 42 (< 0.1%). For exposure groups studied, the median minimum SAS in each time interval was 3.

Female patients had higher minimum SAS than male patients (OR 1.41, CrI 1.23 to 1.62, Figure 3 & S9). Using the observed 19% probability of deep sedation (minimum SAS 2 or lower) in the average male patient, this amounted to an 4.5% decrease (CrI 3.0 to 6.4) in the probability of deep sedation for a female patient. Minimum SAS was lower in Black compared to White patients (OR 0.78, CrI 0.65 to 0.95). Using the observed 19% probability of deep sedation for an average White patient, this translated to a 4.1% absolute increase (CrI 0.8 to 7.5) in the probability of deep sedation for a Black patient. Asian and Hispanic patients had similar minimum SAS to White patients.

**Figure 3:**
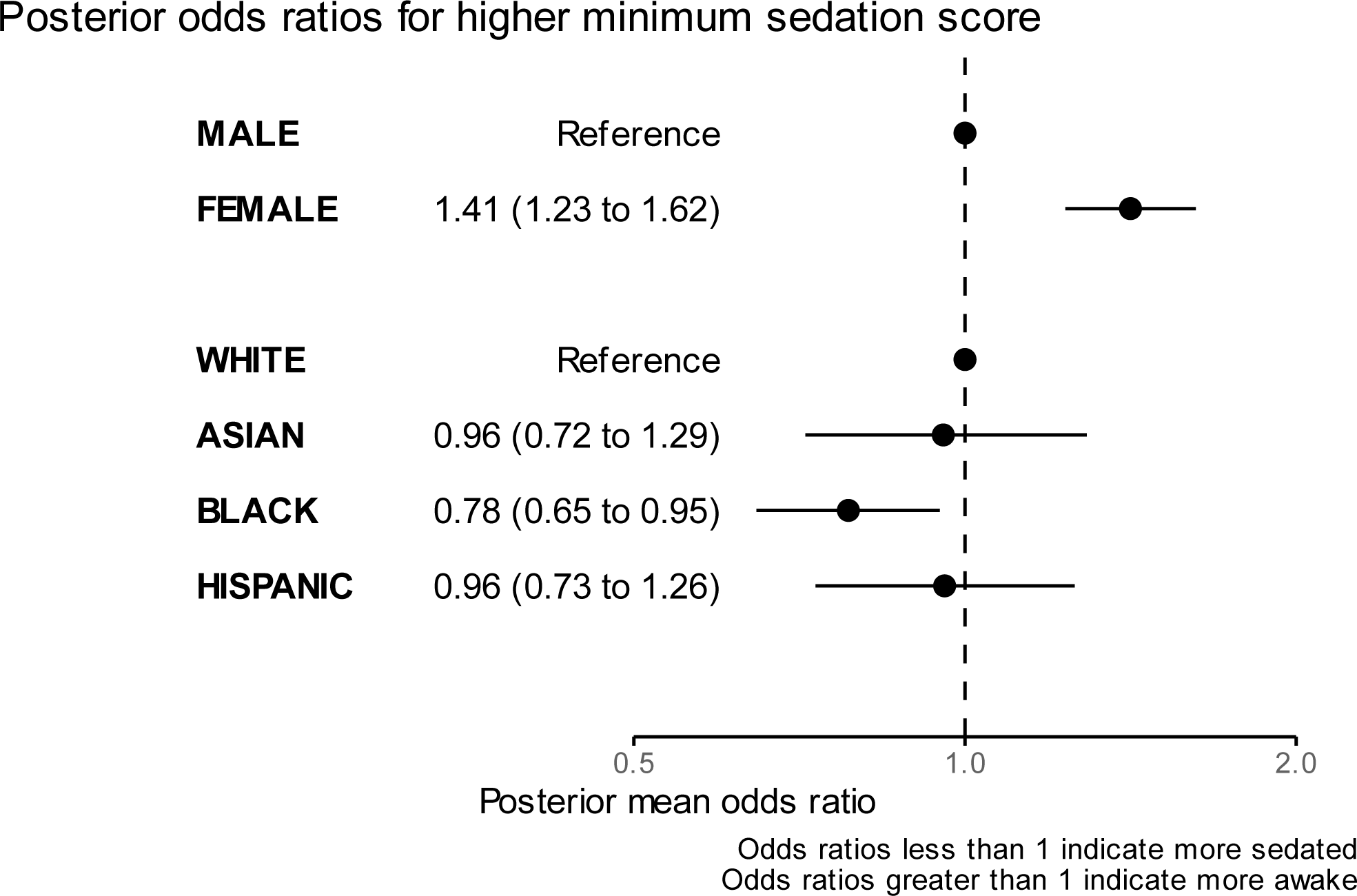
Minimum SAS by Sex and Race and Ethnicity. This figure shows the posterior odds ratios for higher minimum sedation score by sex (top section) and race and ethnicity (bottom section), after adjusting for baseline and time-varying variables. Odds ratios less than 1 correspond to an association with lower minimum SAS, and odds ratios greater than 1 correspond to an association with higher minimum SAS.

## Discussion

### Main findings

In this retrospective single-center cohort study of 6,764 adults invasively ventilated for at least 24 hours, we used multilevel Bayesian proportional odds modeling to show that the use of weight-based propofol, benzodiazepines, and dexmedetomidine, and the levels of sedation attained using these sedatives, varied by sex and race and ethnicity. Female patients compared to male patients received less propofol and were less sedated. Results for Asian patients were more uncertain, but Asian patients compared to White patients probably received less benzodiazepines, propofol, and dexmedetomidine, with similar attained sedation levels. Black patients compared to White patients received less benzodiazepines and propofol, yet were more sedated. Hispanic patients compared to White patients probably received less benzodiazepines, but were similar with respect to propofol, dexmedetomidine, and sedation level.

### Findings in context of prior research

In many prior studies of sedation use, no differences in sedation management by sex were investigated or reported.(4,5,30–32) A recent secondary analysis of control group patients from a multi-centre randomized trial focused on ARDS was designed to investigate differences in sedation levels according to ethnicity, but it also showed that the odds of a male compared to female patient being deeply sedated was 1.27, with wide confidence intervals.(13) In our study, we found a similar point estimate for the odds of male compared to female patients being deeply sedated (1.41, CrI 1.23 to 1.62). We also showed that propofol use was lower in female patients. Taken together, these findings suggest that excess propofol in male patients could be overtreatment.

In comparison to existing research on the association between race and ethnicity and the use of invasive ventilation, our findings have both differences and similarities. A secondary analysis of a multicentre trial of patients with ARDS showed that Hispanic patients had 5 times higher odds of deep sedation than non-Hispanic White patients.(13) A multicentre registry of COVID-19 ICU patients found that Hispanic patients were more likely to receive benzodiazepines than non-Hispanic White or non-Hispanic Black patients.(14) These differ from our finding that Hispanic patients most likely received less benzodiazepine than White patients, similar benzodiazepine to Black patients, and had similar sedation scores to White patients. These differences could be due to the lack of COVID-19 patients in our cohort, differences in unmeasured confounders between our single-center study and the two multicentre studies, or differences due to increased heterogeneity in our patient population compared to populations with COVID-19 or ARDS. By contrast, our results are more consistent with a multi-centre retrospective study showing that Black compared to White patients spent more time deeply sedated in the first 48 hours of ICU admission.(15) This study focused on a similar population of mixed medical-surgical ICU patients with at least 24 hours of invasive ventilation. Our findings and these three studies demonstrate that sex and race and ethnicity can be associated with sedation management, even if the associations vary by context.

### Potential explanations and implications of the findings

There are several potential explanations for the differences in sedation practices that we found. There may be biological differences in pharmacokinetics or pharmacodynamics of sedatives according to patient sex or race and ethnicity.(33) However, race and ethnicity are socially constructed entities with minimal biological correlation; differences here are unlikely to relate to intrinsic pharmacology.(34) Instead, differences are more likely to relate to unmeasured additional factors associated with exposures, such as the nature of co-administered medications and their interactions, or differences in renal, hepatic, and neurologic function due to critical illness or baseline comorbidities. Further confounding could relate to differences in underlying diseases, patient agitation not reflected in the minimum SAS, or nurse staffing.(35)

Implicit bias is also a potential explanation for differences in care according to sex or race and ethnicity.(36,37) Acknowledging the limited external validity of a single-center study, we showed that privileged categories (male, white) generally received sedation most frequently and in the highest doses. This may be an example where patients with relative social or cultural privilege received overtreatment.(12) In the context of sedation, overtreatment is probably detrimental to patient outcomes.(4)

Our findings for Black patients were also consistent with overtreatment. However, we found that Black patients compared to White patients received less propofol and benzodiazepines, despite being more sedated. Given the relatively small proportion of Black patients in our sample, clinical practice may have been unintentionally optimized to treat patients with the diagnoses, medications, and comorbidities associated with White patients at this centre. The findings could be explained by inadequate adaptation of sedation practices to the diagnoses, medications, and comorbidities associated with Black patients at this centre.

Although many prior observational studies have attempted to infer an association between sedation use and mortality or duration of ventilation, we have not done so, because we were not convinced that sufficient control of unmeasured confounding could be achieved in this data. This makes the implications of our findings more difficult to interpret. However, guidelines recommend using the minimum amount of sedation required to achieve safe clinical care, suggesting that we have identified an opportunity to provide less propofol to male patients and target lighter sedation in Black patients.(6,38)

### Limitations

This study has important limitations. It was a single-center study with uncertain external validity. We chose not to attempt analyses linking differences in sedation to outcomes such as mortality due to unmeasured confounding between sedation use and mortality. There were also important unmeasured confounders between race and ethnicity and sedation practices, most notably whether the clinical goal was to provide deep sedation or minimal sedation. We did not include information on parameters such as glomerular filtration rate or drug interactions that might alter clearance. Some confounders that we included were measured in imperfect ways, such as insurance status as a proxy for socioeconomic status, or International Classification of Disease codes in discharge summaries to identify comorbidities. We only had data on female and male sex, with no other sexes charted. A further limitation is that race and ethnicity are distinct entities, while the data used for this study combined them into one field. The race and ethnicity categories are broad and contain important heterogeneity not captured in this analysis. The results themselves may be unique to the social, cultural, and healthcare systems of this particular centre in the United States of America, and may not generalize to other contexts.

## Conclusion

Among patients invasively ventilated for at least 24 hours, intravenous sedation and attained sedation levels varied by sex and race and ethnicity. Adherence to sedation guidelines may improve equity in sedation management for critically ill patients.

## Supporting information

Electronic supplement

## Data Availability

All data are available online at https://physionet.org/content/mimiciv/.

