## Supplementary material for "Association between sex and race and ethnicity and intravenous sedation use in patients receiving invasive ventilation": Electronic supplement

Sarah Walker et. al

2024-04-04

#### Strengthening the Reporting of Observational Studies in Epidemiology checklist

| | Item No | Recommendation | Page No |
|---|---|---|---|
| **Title and abstract** | 1 | (*a*) Indicate the study's design with a commonly used term in the title or the abstract | 1 |
| | | (*b*) Provide in the abstract an informative and balanced summary of what was done and what was found | 5 |
| **Introduction** | | | |
| Background/rationale | 2 | Explain the scientific background and rationale for the investigation being reported | 6 |
| Objectives | 3 | State specific objectives, including any prespecified hypotheses | 6 |
| **Methods** | | | |
| Study design | 4 | Present key elements of study design early in the paper | 8 |
| Setting | 5 | Describe the setting, locations, and relevant dates, including periods of recruitment, exposure, follow-up, and data collection | 7 |
| Participants | 6 | (*a*) Give the eligibility criteria, and the sources and methods of selection of participants. Describe methods of follow-up | 8-11 |
| | | (*b*) For matched studies, give matching criteria and number of exposed and unexposed | |
| Variables | 7 | Clearly define all outcomes, exposures, predictors, potential confounders, and effect modifiers. Give diagnostic criteria, if applicable | 8-11 |
| Data sources/ measurement | 8* | For each variable of interest, give sources of data and details of methods of assessment (measurement). Describe comparability of assessment methods if there is more than one group | 7-11 |
| Bias | 9 | Describe any efforts to address potential sources of bias | 8-11 |
| Study size | 10 | Explain how the study size was arrived at | N/A |
| Quantitative variables | 11 | Explain how quantitative variables were handled in the analyses. If applicable, describe which groupings were chosen and why | 8-11 |
| Statistical methods | 12 | (*a*) Describe all statistical methods, including those used to control for confounding | 9-10 |
| | | (*b*) Describe any methods used to examine subgroups and interactions | |
| | | (*c*) Explain how missing data were addressed | |
| | | (*d*) If applicable, explain how loss to follow-up was addressed | |
| | | (*e*) Describe any sensitivity analyses | |
| **Results** | | | |
| Participants | 13* | (a) Report numbers of individuals at each stage of study—eg numbers potentially eligible, examined for eligibility, confirmed eligible, included in the study, completing follow-up, and analysed | Figure S2 |
| | | (b) Give reasons for non-participation at each stage | |
| | | (c) Consider use of a flow diagram | |
| Descriptive data | 14* | (a) Give characteristics of study participants (eg demographic, clinical, social) and information on exposures and potential confounders | 11, Table 1 |
| | | (b) Indicate number of participants with missing data for each variable of interest | 12, supplement |
| | | (c) Summarise follow-up time (eg, average and total amount) | |
| Outcome data | 15* | Report numbers of outcome events or summary measures over time | Table 2, p11-15 |

| | | | |
|---|---|---|---|
| Main results | 16 | (*a*) Give unadjusted estimates and, if applicable, confounder-adjusted estimates and their precision (eg, 95% confidence interval). Make clear which confounders were adjusted for and why they were included<br>(*b*) Report category boundaries when continuous variables were categorized<br>(*c*) If relevant, consider translating estimates of relative risk into absolute risk for a meaningful time period | p11-14 |
| Other analyses | 17 | Report other analyses done—eg analyses of subgroups and interactions, and sensitivity analyses | P15 |
| **Discussion** | | | |
| Key results | 18 | Summarise key results with reference to study objectives | 16 |
| Limitations | 19 | Discuss limitations of the study, taking into account sources of potential bias or imprecision. Discuss both direction and magnitude of any potential bias | 19-20 |
| Interpretation | 20 | Give a cautious overall interpretation of results considering objectives, limitations, multiplicity of analyses, results from similar studies, and other relevant evidence | 16,20 |
| Generalisability | 21 | Discuss the generalisability (external validity) of the study results | 16-20 |
| **Other information** | | | |
| Funding | 22 | Give the source of funding and the role of the funders for the present study and, if applicable, for the original study on which the present article is based | 4 |

### Data Extraction & Processing

#### Extracting Patient Eligibility

We are using the MIMIC-IV database, so all referenced datasets are from that database.

First, we connected the icu.icustays dataset (which details each unique patient stay in the ICU) with the hosp.patients dataset and the hosp.admissions dataset (the latter two containing information about the patient and their general hospital stay). From there, we extracted sex and race and ethnicity, if recorded, for each patient's unique stay in the ICU. Race and ethnicity were grouped into the categories Asian, Black, Hispanic, and white. We excluded the category Indigenous due to a lack of patients (64 total), as well as the categories Multiple and Other.

Using the derived ventilation table, we calculated whether each ICU stay was eligible based on having received invasive ventilation for at least 24 hours. We did this by extracting start and end times for when patients received invasive ventilation (excluding tracheostomies). If a patient had recorded events of invasive ventilation that started and stopped within 48 hours of each other, we connected the two events as if there was no stop in between. Therefore, we calculated the duration of each invasive ventilation period as the time in minutes from start to end time where the next event's start time was the first of:

A) A period of invasive ventilation, but greater than 48 hours after the previous end time, or
B) A period of non-invasive ventilation or a tracheostomy, or
C) Did not exist; there was no next recorded ventilation event.

We also excluded events that were recorded to start more than 12 hours before the patient's ICU admission time, as recorded in the icu.icustays dataset. From those remaining, we took the first period of invasive ventilation that lasted for more than 24 hours.

Then, for each patient, we excluded all those who had received a tracheostomy prior to their period of invasive ventilation. We did this by calculating tracheostomy periods using the same metric as with the invasive ventilation periods and comparing the patient's tracheostomy end times to that of the same patient's invasive ventilation start time.

We also used the death date information from the icu.icustays to exclude patients whose death dates were earlier than the period of invasive ventilation; these cases were presumed to be from brain death and/or organ donors. We also recorded whether each patient received invasive ventilation at all, regardless of whether they received it for more than 24 hours.

#### Extracting Baseline Covariates

Using only the ICU stays that were marked as eligible in the previous table, we extracted sex, race and ethnicity, time from invasive ventilation period to death (if occurred), and duration of their period of invasive ventilation.

Using the hosp.patients dataset, we extracted each patient's age, and approximate year of hospital admission. Using the hosp.admissions dataset, we extracted their hospital LOS in days, their English proficiency (proficient or uncertain) and their insurance status. Using the icu.icustays dataset, we extracted the ICU LOS in days and the ICU type upon admission (cardiac, neuro-trauma, medical-surgical). We extracted the patient's height in cm from the derived table height, and their weight in kg from the derived table first_day_weight. Finally, we extracted each patient's past history with dementia, substance abuse, and traumatic brain injury from the hosp.diagnoses_icd dataset using ICD-9 and ICD-10 codes as binary variable (1 as occurred / 0 as did not).

*Table S1: ICD Codes Used*

| Item | ICD Code(s) | ICD Version |
|---|---|---|
| Dementia | F0-3 | 10 |
| | 290 | 9 |
| Traumatic Brain Injury | S020-S021 | 10 |
| | S0281-S0283 | |
| | S0291 | |
| | S0402-S0404 | |
| | S071 | |
| | T744 | |
| | 801-804 | 9 |
| | 850-854 | |
| | 951-953 | |
| | 95901 | |
| | 99555 | |
| Substance Use Disorder | 291-292 | 9 |
| | 3050 | |
| | 3575 | |
| | 3030 | |
| | 3039 | |
| | 4255 | |
| | 5353 | |
| | 5710-5713 | |
| | 6554 | |
| | 76071 | |
| | 9800-9801 | |
| | 64830-64834 | |
| | 3040-3049 | |
| | 3052-3057 | |
| | 3059 | |
| | 9650 | |
| | 9685 | |
| | E8600-E8602 | 10 |
| | E8609 | |
| | G621 | |
| | G312 | |
| | I426 | |
| | X45 | |
| | X65 | |
| | K292 | |
| | K700-K704 | |
| | K709 | |
| | K852 | |
| | K860 | |
| | Q860 | |
| | P043 | |
| | Y15 | |
| | F10-F19 | |
| | V6542 | |
| | E0385 | |
| | E9350 | |
| | E9396 | |
| | E8500 | |
| | E8541 | |

#### Extracting Time-Varying Variables

##### Drug Dosages

First, we extracted all recorded dosages of intravenous opioids, vasopressors/inotropes, propofol, benzodiazepines, and neuromuscular blockers for each 4 hour time interval using the icu.inputevents dataset.

We did this by recording the following information for each recorded instance of the above drugs:

- the minute offset to the end of the patient receiving invasive ventilation (with 0 being the patient's ICU admission time),
- the time interval at which the drug was first administered (with time interval 0 as 24 hours after beginning invasive ventilation),
- the minute offset at which the drug was first administered (with minute 0 as their ICU admission time),
- the time interval at which the drug stopped being administered (with time interval 0 as 24 hours after beginning invasive ventilation),
- the minute offset at which the drug stopped being administered (with minute 0 as their ICU admission time),
- the total quantity administered over the whole duration (in mcg, unless the drug is vasopressin which is stored in units),
- and the name of the drug administered.

In order to calculate the various offsets, we used the datasets icu.icustays (for ICU admission time), the previously calculated baseline table (for ventilation duration), and the previously calculated eligibility table (for eligible patients, and the minute offset of first receiving invasive ventilation). This information was calculated with the restriction that the drug was administered no earlier than their first moment receiving invasive ventilation.

In a recursive manner, we averaged the total dosage administered over as many time intervals as the total duration lasted. Calculating the dosage for time intervals was done proportionally; the average amount of dose administered per minute was multiplied by the length of the new interval. For most cases, this amounted to 4 hours, but the last time interval of that event would reflect however many minutes into the last interval the drug was administered instead of the full 4 hours.

Then, we converted the neuromuscular blockers into a binary variable (received / did not receive) and excluded the recorded events that happened after the time interval where the patient was taken off invasive ventilation or were past their first 7 days on invasive ventilation. We also summed up the total amount of propofol and benzodiazepines administered in the first 24 hours in mcg.

##### "Maximum" Covariates

We kept the maximum values of the patient's respiratory rate and FiO2. This was extracted from the icu.chartevents dataset. We restricted respiratory rate values to be between 0 and 70 (exclusive). Similarly, we excluded events that happened after being taken off invasive ventilation, after the patient's first 7 days on invasive ventilation, or occurred before entering their period of invasive ventilation.

##### "Minimum Covariates"

We kept the minimum values of the patient's SpO2 values. This was extracted from the icu.chartevents dataset. We restricted SpO2 values to be between [0, 100]. Similarly, we excluded events that happened after being taken off invasive ventilation, after the patient's first 7 days on invasive ventilation, or occurred before entering their period of invasive ventilation.

##### Table S2: Medications Included

| Group | Medications | MIMIC-IV Itemid(s) |
|---|---|---|
| Opioids | Morphine, Hydromorphone, Fentanyl, Meperidine | 225154; 221833; 221744, 225942, 225972; 225973 |
| Vasopressors | Norepinephrine, Epinephrine, Dopamine, Vasopressin, | 221906, 221289, 221662, 222315 |
| Propofol | Propofol | 222168 |
| Benzodiazepines | Lorazepam, Midazolam, Diazepam | 221385, 221668, 221623 |
| Neuromuscular blockers | Cisatracurium, Vecuronium, Rocuronium | 221555, 222062, 229233 |
| Antipsychotics | Haloperidol | 221824 |
| Dexmedetomidine | Dexmedetomidine | 225150, 229420 |

##### Standardized sedation scales

As outlined in the Variables section of the Methods in the main manuscript, we included standardized sedation scale assessment as a time-varying covariate. MIMIC-IV has both Richmond Agitation and Sedation Scale (RASS) values and Riker Sedation Agitation Scale (SAS) scores. We combined them using a previously established conversion from RASS to SAS.[1] The conjoined sedation/agitation scores were divided into five categories: scores of 1, 2, 3, 4, and 5-7. This was due to few patients receiving SAS scores of 5 or more.

#### Processing for Analysis

##### Joining Baseline and Time-Varying Covariates and Outcomes

For each outcome drug (propofol, benzodiazepines, and dexmedetomidine), we perform the following steps. From the extracted time-varying covariate table, we keep only rows that correspond to the current outcome drug with a time interval greater than or equal to 0 (greater than or equal to 24 hours after receiving invasive ventilation). Then, we convert the table to wide form and merge it with the extracted baseline covariate table on the patient's ICU stay id.

We split up categorical variables in the extracted baseline covariates table into binary ones. For example, the race/ethnicity column was converted into three binary columns (Asian, Black, and Hispanic; if all three columns were 0 then that indicates the patient belongs to the white category). ICU type was converted into the binary columns cardiac and medical-surgical, with the last group being neuro-trauma. Finally, insurance was split into two binary columns: Medicare and Medicaid, with the last group representing other forms of insurance.

Then, using the extracted time-varying covariates table again, we append columns for the other variables. They are taken from the time interval immediately preceding that of the drug dose; for example, if propofol was administered at time interval 1, then the other time-varying covariates would have been recorded in the time interval 0.

The variable FiO2 corresponding to when an outcome drug is administered at time 0 is an exception. We assume that these values will not have substantially changed if changed at all between measurements. As such, their values are taken from the most recent time interval between -6 (24 hours before, right when first receiving invasive ventilation) and -1 (4 hours before). This is to reduce the quantity of missing data. Missing data for these variables at a different time interval are dealt with through the usual imputation method.

We also converted the dosages of vasopressors/inotropes and opioids into base units of norepinephrine. The dosage administered of each other outcome drug in the previous time interval was also included; for example, benzodiazepine and dexmedetomidine were predictors for propofol, but the dose of propofol administered in the previous time interval was not.

In this step, the benzodiazepines lorazepam, midazolam, and diazepam were also combined into one unit (base unit being lorazepam) by dividing the diazepam dosage by 5 and midazolam by 2.

#### Transformation, Centering, and Scaling

The following processes were applied for each outcome drug. The continuous variable of respiratory rate was transformed using the natural logarithm function. The variables SpO2, FiO2, and minimum Riker SAS values were treated as continuous interval variables and transformed with the logit function. To facilitate use of the logit function, which maps the (0,1) interval onto the real line, we converted the observed intervals to the (0,1) interval by centering and scaling. Any values that produced an invalid number from the transformation were imputed the same way as if it had been missing from the start, because those values were outside the range of plausible values for that variable.

The drug dose, whose units were previously (mcg or units)/4 hours, was divided by the patient's weight to get units as (mcg or units)/kg/4 hours. Then, quartiles were computed for non-zero drug dosages. After imputation, each drug dose quantity with the above units was categorized as either 0 (no drug administered) or by the previously computed quartiles.

#### Imputing Missing Data

Multiple imputation by chained equations with 10 datasets was applied to missing time-varying variables at the time interval 0. Those included were respiratory rate, FiO2, SpO2, and the primary outcomes of propofol and benzodiazepines for patients who were missing a recorded weight. This was later converted into clinically relevant units.

Afterwards, missing values were imputed by forward fill grouped by patient's ICU stay id and ordered by time interval.

#### Final Adjustments

Patient age was divided into four categories, based on quartiles. The categories were less than 55, between 55 and 65, between 65 and 75, and greater than 75. The variable time since initiation was grouped into three categories: in the first day, between days 2-3, and between days 4-7. The minimum sedation/agitation score was converted to categories of unarousable (Riker SAS score of 1), very sedated (Riker SAS score of 2), sedated (Riker SAS score of 3), calm (Riker SAS score of 4), and agitated (Riker SAS scores 5-7).

Propofol doses were converted to units of 10mcg/kg/min. Benzodiazepine doses were converted to units of 0.1mg/kg/hr. Dexmedetomidine doses were converted to units of 0.2mcg/kg/hr. Vasopressor doses (in norepinephrine equivalents) were converted to units of 0.1mcg/kg/min. Opioid doses (in morphine equivalents) were converted to units of 1mg/kg/hr. A cap for doses of vasopressors and opioids was imposed, with values above the cap being set to 1mcg/kg/min and above 1mg/kg/hr respectively.

#### Analysis

Proportional odds models grouping by patient stay were run on the Niagara computing cluster. This was done with the brms package in R 4.2.1, with 4 chains, 80 cores, and 20 threads. The first 5 of the 10 imputed datasets were used with the brm_multiple function. The model took a significant amount of time and computational resources, which is part of the reason we did not use more imputations.

##### Sensitivity analysis

The sensitivity analysis considering patients who were previously excluded due to race or ethnicity had a total of 8,933 patients (Figure S3). They were 42% female (3,758). Asian patients comprised 3% (236) of the total, Black patients 9% (806), Hispanic patients 3% (300), white patients 61% (5,422), and those classified as "Other" 24% (2,169).

### Additional tables and figures

#### Table S3: Missing data at baseline

| | TOTAL |
|---|---|
| Age | 0 (0.0) |
| Weight | 278 (0.04) |
| Respiratory rate | 329 (0.05) |
| FiO2 | 117 (0.02) |
| SpO2 | 335 (0.05) |
| Minimum SAS | 1407 (0.21) |
| Dementia | 0 (0.0) |
| TBI | 0 (0.0) |
| Substance use disorder | 0 (0.0) |
| English | 0 (0.0) |
| Insurance | 0 (0.0) |
| Opioids | 0 (0.0) |
| Vasopressors | 0 (0.0) |
| Neuromuscular blockers | 0 (0.0) |

#### Table S4: Time-varying covariates by patient sex

| | TOTAL | FEMALE | MALE |
|---|---|---|---|
| **TOTAL (%)** | 6764 | 2924 (43) | 3840 (57) |
| **Resp. Rate (IQR)** | 22 (18, 26) | 22 (18, 26) | 22 (18, 26) |
| **FiO2 (IQR)** | 40 (40, 50) | 40 (40, 50) | 40 (40, 50) |
| **SpO2 (IQR)** | 97 (95, 99) | 97 (95, 99) | 97 (95, 99) |
| **Sedation/Agitation (IQR)** | | | |
| Min Riker | 3 (3, 4) | 3 (3, 4) | 3 (3, 4) |
| Max Riker | 3 (3, 4) | 3 (3, 4) | 3 (3, 4) |
| **Vasopressors *** | | | |
| # received (%) | 1922 (28) | 794 (27) | 1128 (29) |
| mcg/kg/min (IQR) | 0.11 (0.04, 0.27) | 0.12 (0.05, 0.27) | 0.11 (0.04, 0.27) |
| **Opioid **** | | | |
| # received (%) | 3850 (57) | 1628 (56) | 2222 (58) |
| mg/kg/hr (IQR) | 0.08 (0.03, 0.16) | 0.09 (0.03, 0.16) | 0.08 (0.03, 0.16) |
| **Neuromuscular blocker (%)** | 354 (5) | 138 (5) | 216 (6) |

#### Table S5: Time-varying covariates by patient race and ethnicity

| | TOTAL | ASIAN | BLACK | HISPANIC | WHITE |
|---|---|---|---|---|---|
| **TOTAL (%)** | 6764 | 236 (3) | 806 (12) | 300 (4) | 5422 (80) |
| **Resp. Rate (IQR)** | 22 (18, 26) | 21 (17, 26) | 22 (18, 26) | 22 (18, 26) | 22 (18, 26) |
| **FiO2 (IQR)** | 40 (40, 50) | 40 (40, 50) | 40 (40, 50) | 40 (40, 50) | 40 (40, 50) |
| **SpO2 (IQR)** | 97 (95, 99) | 98 (95, 99) | 98 (96, 100) | 98 (95, 99) | 97 (94, 99) |
| **Sedation/Agitation (IQR)** | | | | | |
| Min Riker | 3 (3, 4) | 3 (2, 4) | 3 (3, 4) | 3 (2, 4) | 3 (3, 4) |
| Max Riker | 3 (3, 4) | 3 (3, 4) | 3 (3, 4) | 3 (2, 4) | 3 (3, 4) |
| **Vasopressors *** | | | | | |
| # received (%) | 1922 (28) | 63 (27) | 210 (26) | 63 (21) | 1586 (29) |
| mcg/kg/min (IQR) | 0.11 (0.04, 0.27) | 0.10 (0.04, 0.35) | 0.10 (0.04, 0.23) | 0.12 (0.05, 0.28) | 0.11 (0.04, 0.27) |
| **Opioid **** | | | | | |
| # received (%) | 3850 (57) | 123 (52) | 417 (52) | 174 (58) | 3136 (58) |
| mg/kg/hr (IQR) | 0.08 (0.03, 0.16) | 0.09 (0.03, 0.19) | 0.08 (0.03, 0.15) | 0.10 (0.04, 0.17) | 0.08 (0.03, 0.16) |
| **Neuromuscular blocker (%)** | 354 (5) | 12 (5) | 37 (5) | 15 (5) | 290 (5) |

* Norepinephrine equivalents
** Morphine equivalents

Figure S1: Conceptual diagram

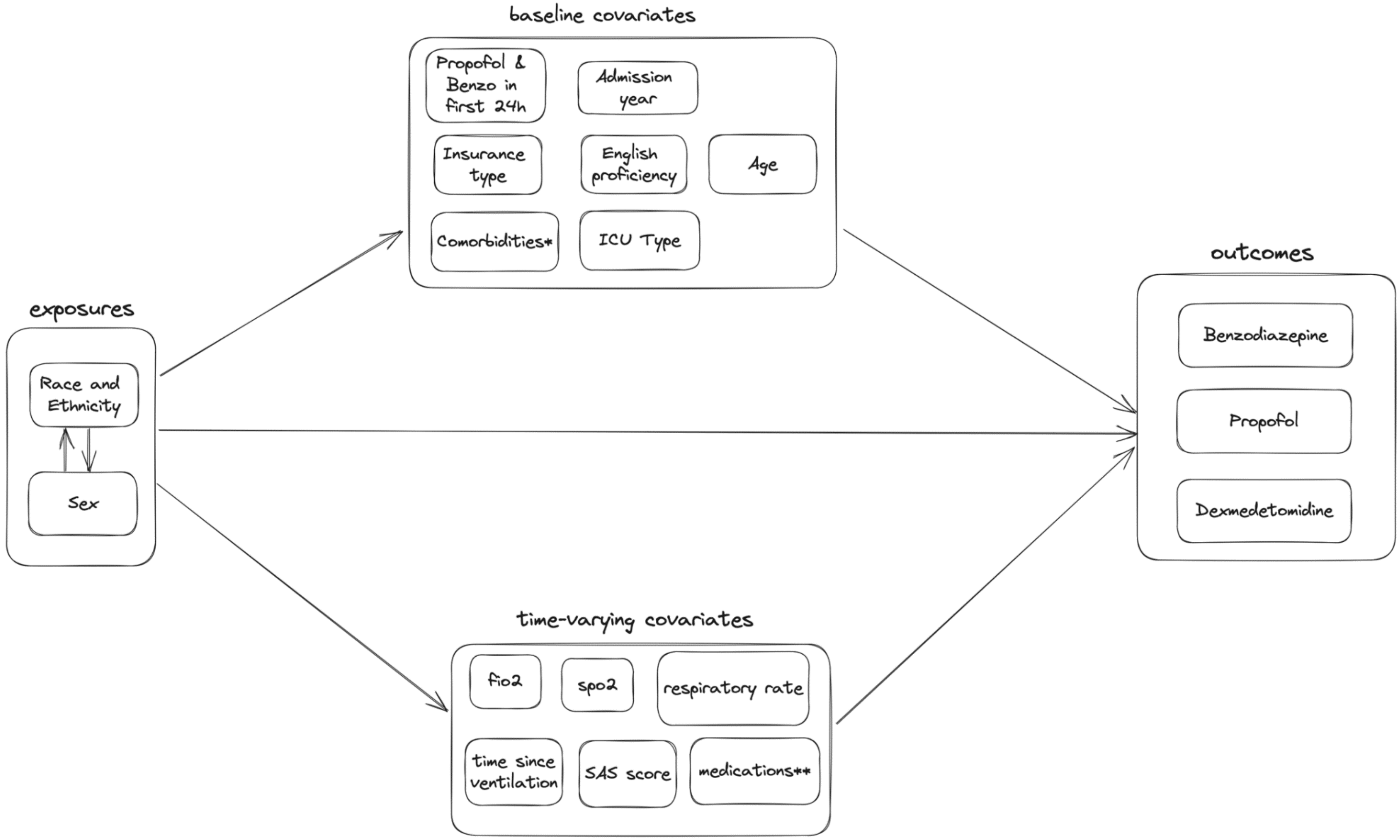

#### Figure S2: Cohort flow diagram for primary cohort

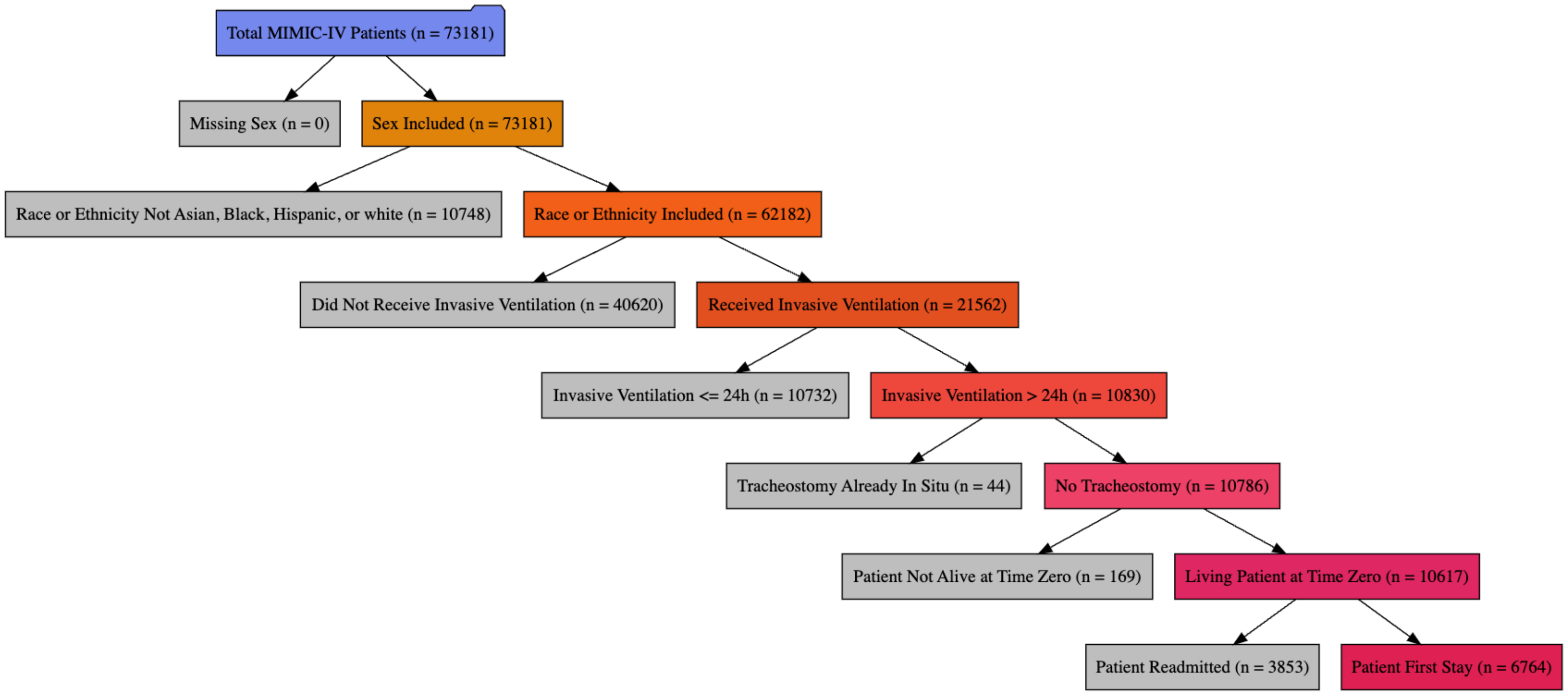

Caption: This figure shows the flow chart for including patients in the study. "Race or ethnicity excluded" corresponds to the patients with "Unknown", "Other", or "Native American / Pacific Islander" noted as their race and ethnicity. MIMIC-IV = Medical Information Mart for Intensive Care version IV.

Figure S3: Cohort flow diagram for sensitivity analysis including patients with race and ethnicity categorized as "Other"

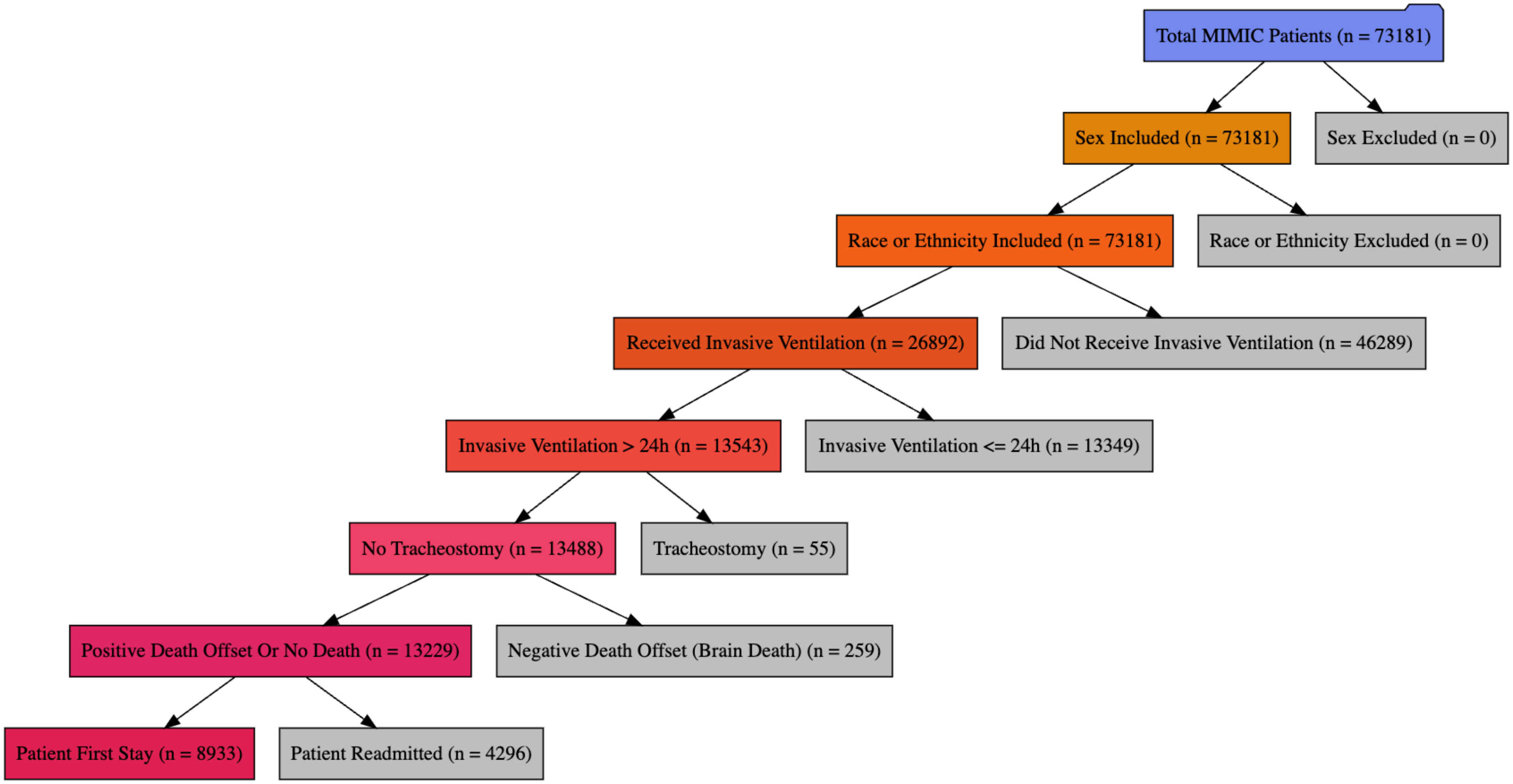

#### Figure S4: Forest plot of odds ratios for all fixed-effect coefficients, benzodiazepine model

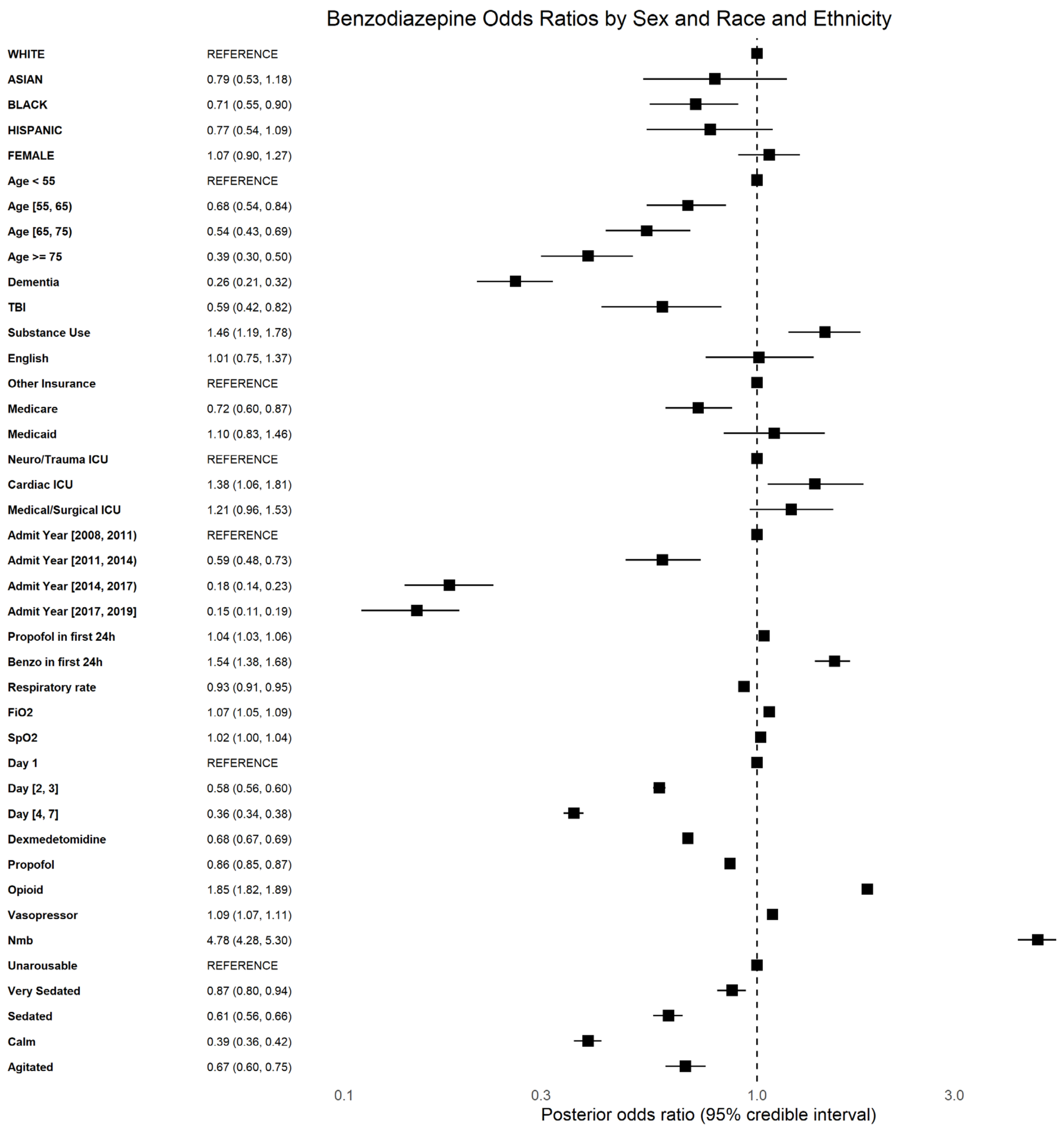

#### Figure S5: Odds ratios by sex and race and ethnicity

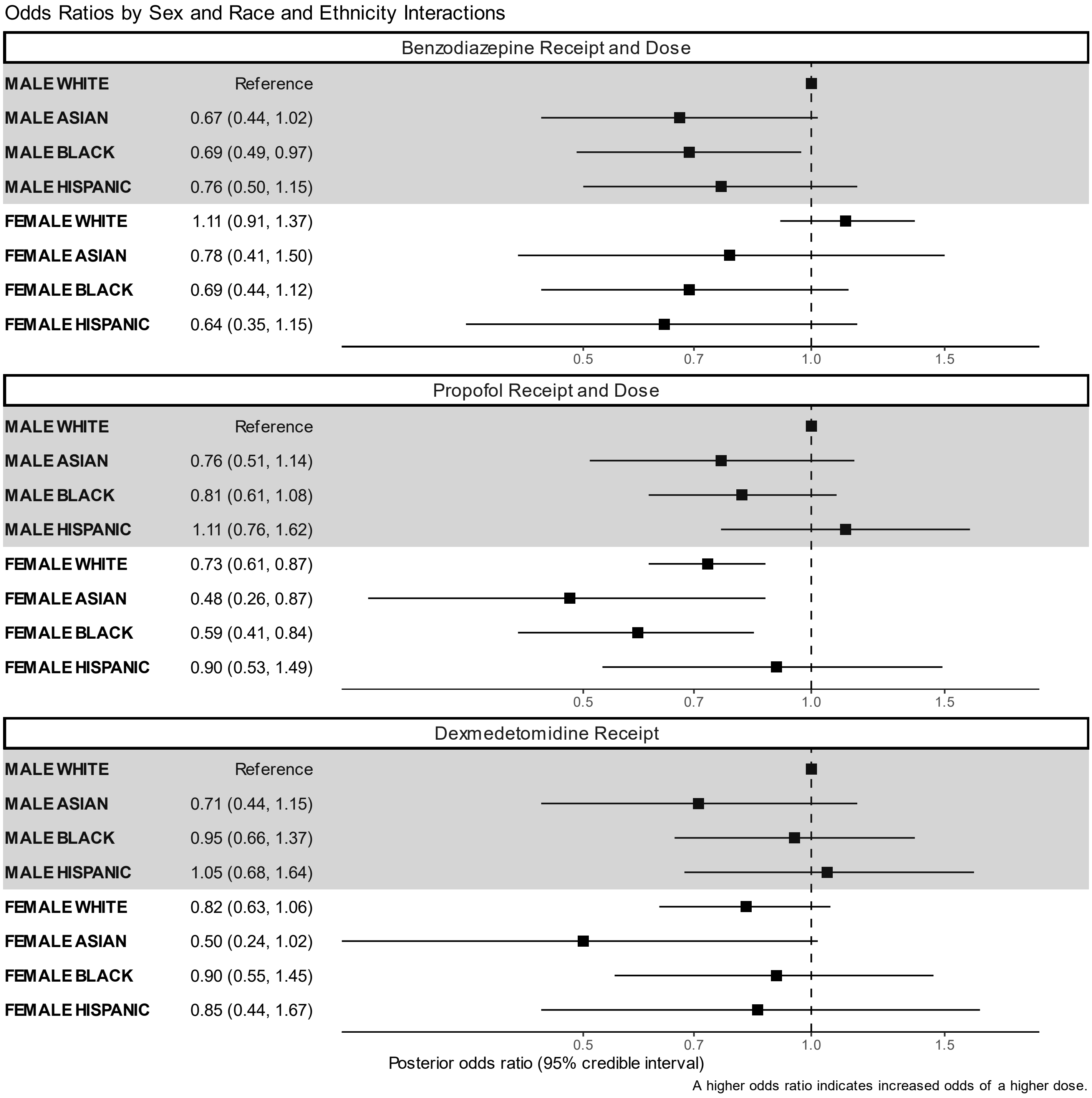

Caption: This figure shows the interactions between sex and race and ethnicity by plotting the odds ratios for receiving a higher dose of each medication according to the sex and race and ethnicity subgroup. Odds ratios less than one correspond to lower doses and lower likelihood of receiving the medication, while odds ratios greater than one correspond to higher doses and higher likelihood of receiving the medication. For dexmedetomidine, the odds ratios reflect only the probability of receiving the medication.

Figure S6: Forest plot of odds ratios for model including “Other” race and ethnicity

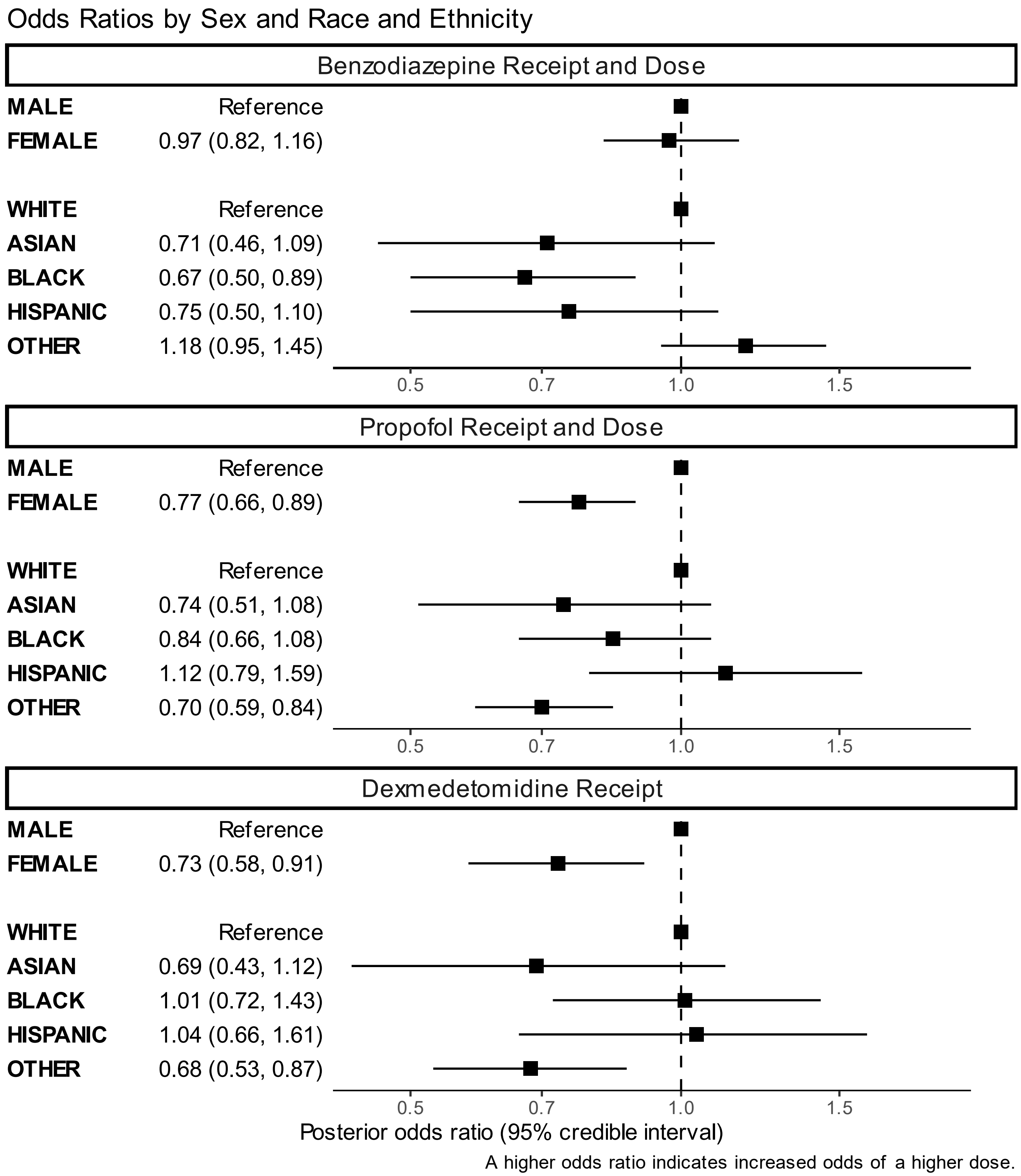

### Figure S7: Forest plot of odds ratios for all fixed-effect coefficients, propofol model

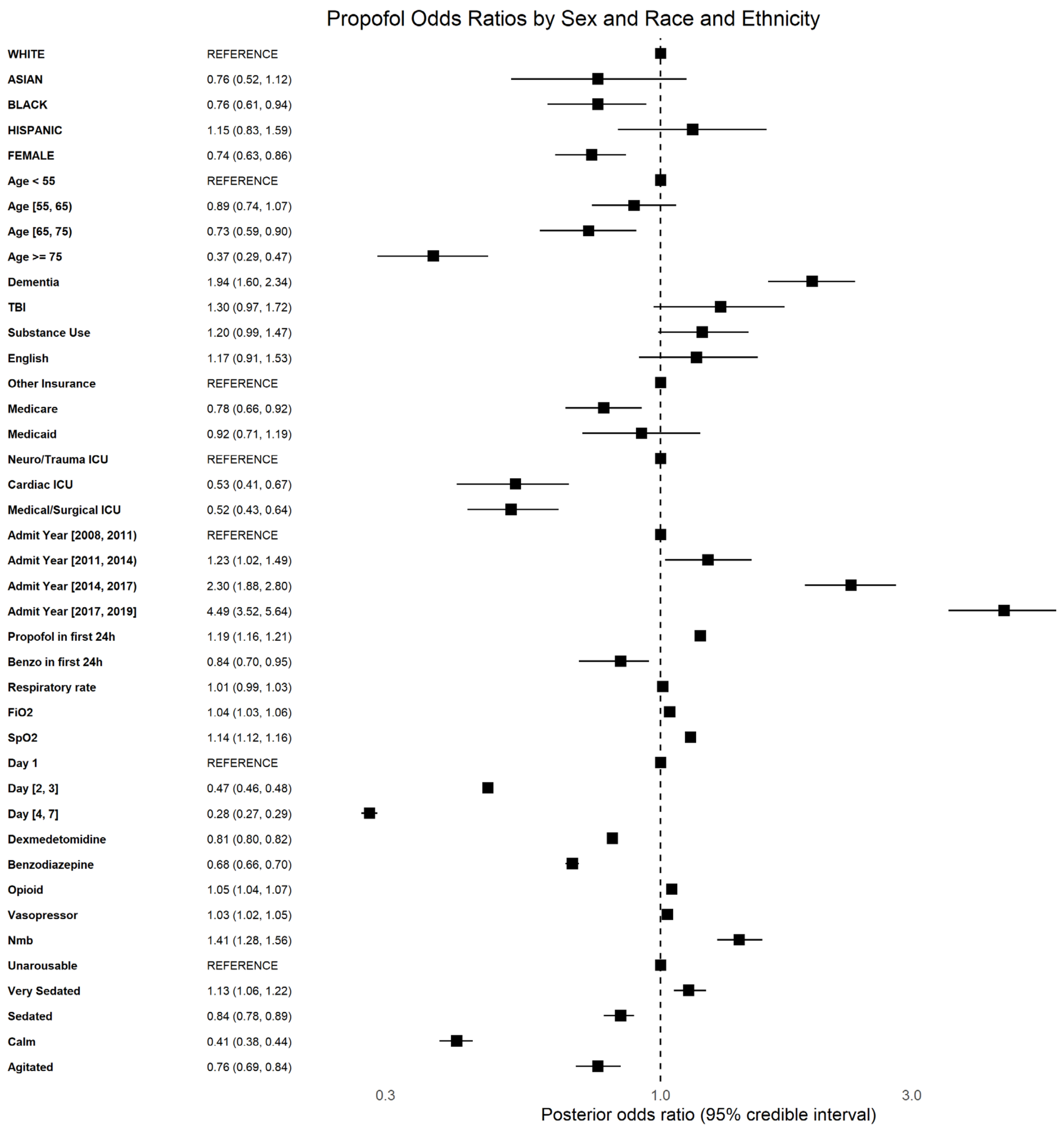

Figure S8: Forest plot of odds ratios for all fixed-effect coefficients, dexmedetomidine model

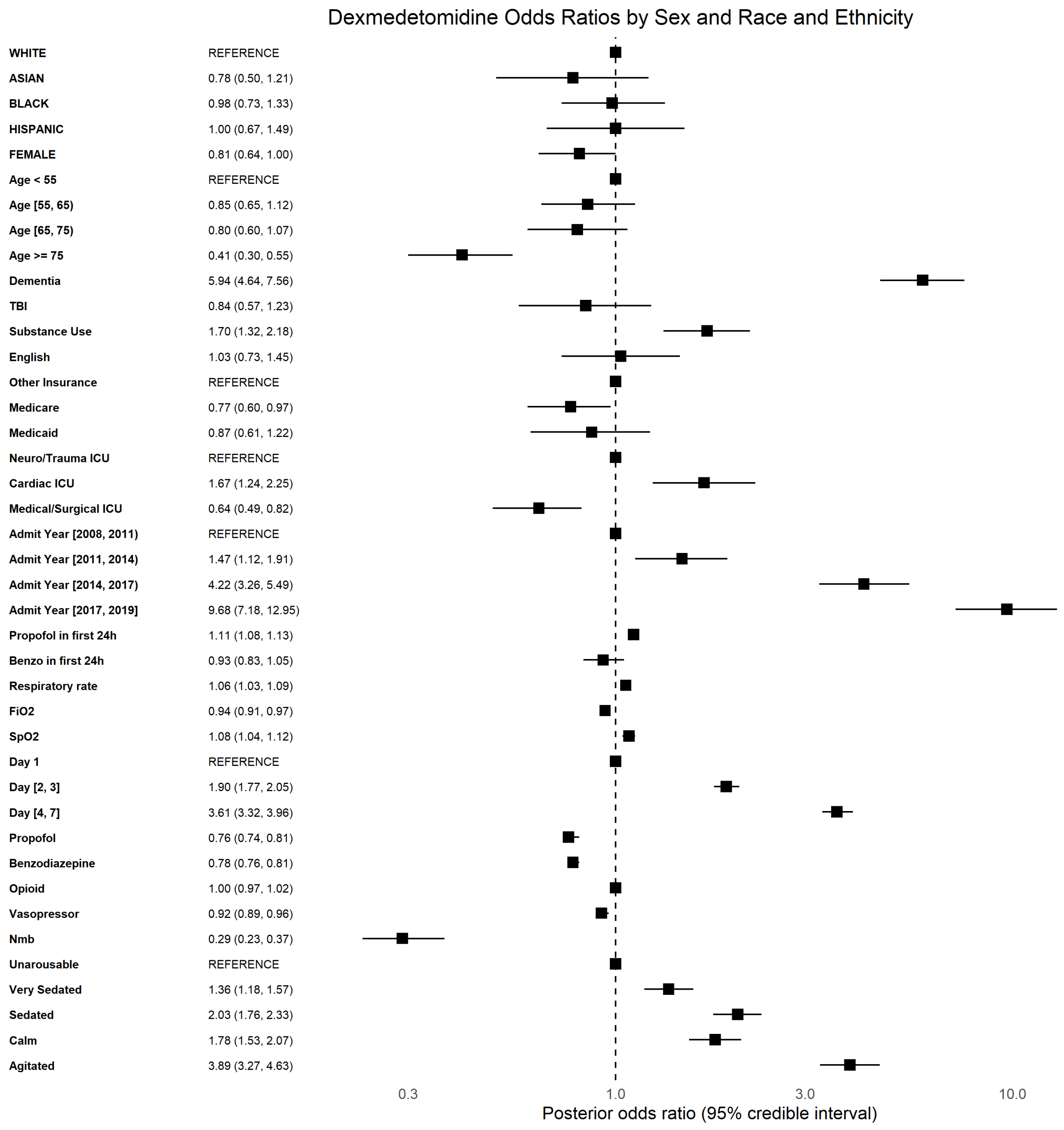

Figure S9: Forest plot of odds ratios for minimum Riker sedation score model

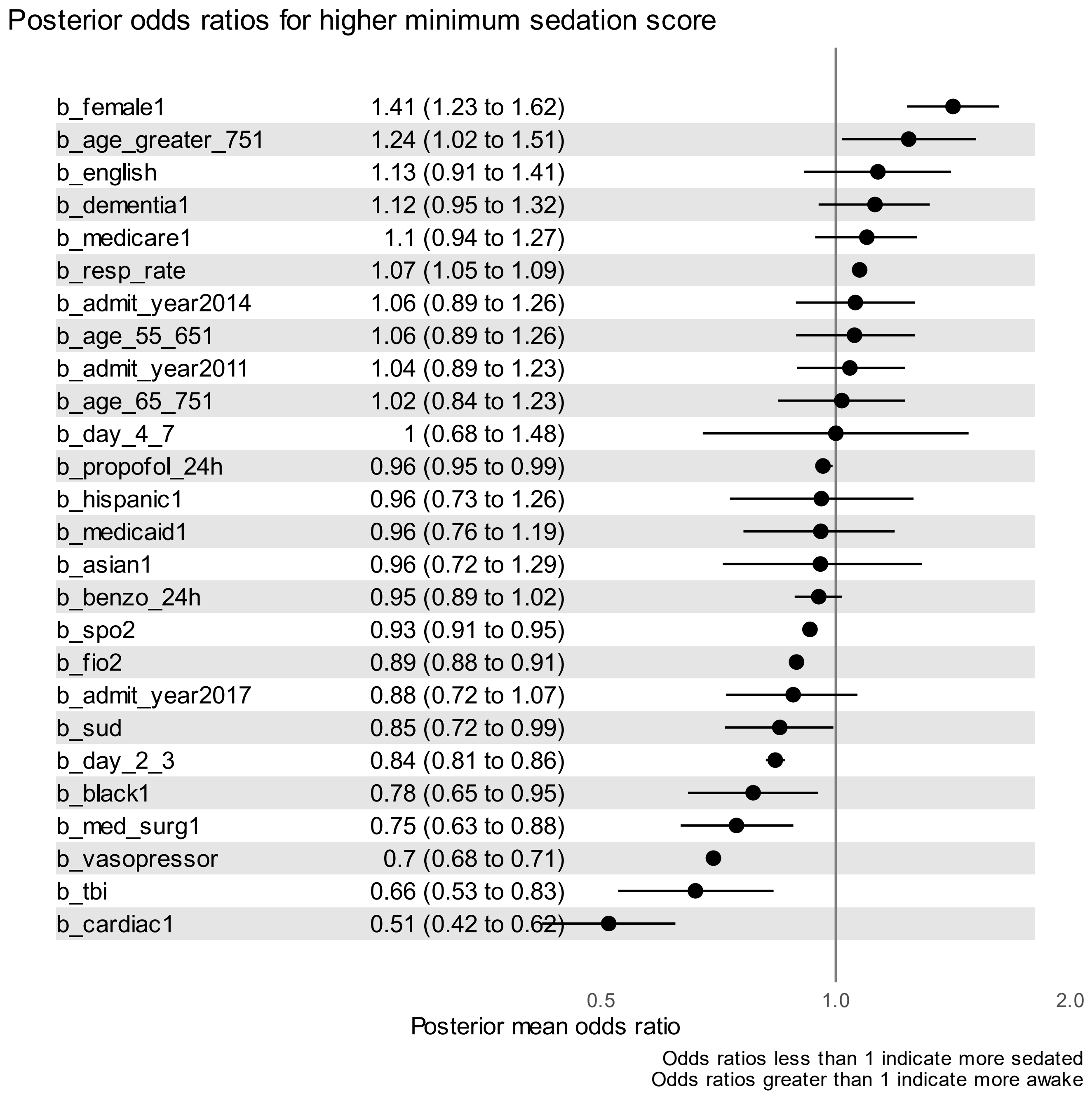

Caption: This figure shows the posterior odds ratios for all of the fixed effect coefficients in the model of minimum SAS in each 4 hour interval. Reference categories vary by coefficient, and are listed here: sex – male, age – less than 55, dementia – no dementia, English proficiency – low English proficiency, admit year – 2019, insurance type – other, day of admission – day 1, average propofol dose in the first 24 hours – 1 mcg/kg/min, race and ethnicity – White, average benzodiazepine dose in the first 24 hours – 0.1mg/kg/hr, substance use disorder (sud) – no substance use disorder, icu type (medical-surgical, cardiac, trauma-neuro) – trauma-neuro, traumatic brain injury (tbi) – no traumatic brain injury
